# Academic achievement in Ugandan children with sickle cell anaemia: A cross-sectional study

**DOI:** 10.1101/2024.07.08.24309901

**Authors:** Shubaya Kasule Naggayi, Paul Bangirana, Robert O. Opoka, Simple Ouma, Betty Nyangoma, Annet Birabwa, Grace Nambatya, Maxencia Kabatabaazi, Ann Jacqueline Nakitende, Dennis Kalibbala, Deogratias Munube, Phillip Kasirye, Ezekiel Mupere, John M. Ssenkusu, Nancy S. Green, Richard Idro

## Abstract

**Objective:** Academic achievement in school-age children is crucial for advancing learning goals. Children with sickle cell anaemia (SCA) in Sub-Saharan Africa may be at risk of disease-associated school difficulties. Limited data exist on the academic achievement of children with SCA in the region. This study aimed to assess academic achievement of children with SCA in Uganda compared to siblings without SCA.

**Design and setting:** A cross-sectional study conducted at Mulago Hospital SCA Clinic in Uganda.

**Participants:** School-going children (6-12 years) with SCA and age-matched sibling controls without SCA.

**Outcome measures:** Academic achievement was tested using the Wide Range Achievement Test, Fourth Edition (WRAT4). Outcome measures were spelling, mathematical computation, word reading, and sentence comprehension by age-normalized Z-scores on the WRAT4 test.

**Results:** Among 68 SCA and 69 control, the mean age (standard deviation) was 9.44 (2.04) and 9.42 (2.02) years and males were 55.9% and 46.4% respectively. Mean haemoglobin was 7.9 (SD 0.89)g/dL in the SCA group versus 12.8 (SD 0.89)g/dL in the controls, (p<0.001). Children with SCA scored lower in spelling, (mean difference [95% confidence interval] - 0.36 [−0.02 to −0.69], *p*=0.04) and mathematical computation, (mean difference [95% confidence interval] −0.51 [−0.17 to −0.85], *p*=0.003) than the controls. In the SCA group, lower scores in spelling correlated with age, while males performed better than females in mathematical computation.

**Conclusion:** School-aged children with SCA are at risk of poor performance in spelling and mathematical computation. Our findings support the need for educational evaluation and possible support, especially in these two areas.

**ARTICLE SUMMARY:** 

**Article focus:** Using a standardized assessment tool, this report provides data on academic achievement in school-age children with sickle cell anaemia (SCA) in Uganda compared to sibling controls.

**Key messages:** School-aged children with SCA may experience academic challenges in key areas of spelling and mathematical computation. These findings suggest a role for educational evaluation and possible support for school-aged children with SCA especially in spelling and mathematics.

**Strengths and limitations of this study:**

- This is one of few studies to investigate academic achievement among children with SCA in sub-Saharan Africa, and the first in East Africa.
- The study used the widely recognised and validated assessment tool, the Wide Range Achievement Test, Fourth Edition (WRAT4), to standardize the measurements and permit regional comparisons.
- Selection of age-matched sibling controls minimised the potential confounding effects of age, socioeconomic status, and environmental factors.
- However, data on school absenteeism, which can affect academic achievement and which is more common in children with SCA, were not collected in this study.

## INTRODUCTION

### Background

Sickle cell anaemia (SCA), the most severe form of sickle cell disease (SCD), is an inherited haematological disorder resulting from the sickle variant in both haemoglobin (Hb) alleles [1]. SCA is most commonly defined as homozygous HbSS, accounting for over 80% of children born with SCD globally [2, 3]. Worldwide, nearly 400,000 children are born annually with SCA, most in sub-Saharan Africa (SSA) [4]. In Uganda, approximately 20,000 children are born with SCD annually, nearly all with SCA [5]. These children face multiple disease-related complications that can impede academic proficiency, including severe anaemia, cerebral infarcts, and illness-related school absences [6, 7].

Academic achievement is crucial in determining a child’s ability to develop into a productive self-sufficient adult [8]. Skills such as reading, writing, and math computation play a key role in literacy, an essential feature in future functioning, e.g. self-management [9, 10]. Academic achievement in children with SCD has been well-studied in high-resource countries [7]. In the United States (U.S.), poor academic achievement has been reported among school-aged children with SCD, especially those aged 6 years or older [7, 11, 12]. In contrast, limited research on this has been conducted in SSA, despite having the largest SCA burden, more frequent severe complications (e.g. cerebral infarcts), adverse environmental conditions (e.g. malaria and other infections), poverty and less access to SCA treatment [13, 14].

Studies in Nigeria have demonstrated slower academic progress and below-average academic achievement scores among children with SCA compared to their healthy peers [15-18]. However, there is a lack of comparative research examining the academic outcomes of children with SCA in diverse geographical and educational settings in SSA outside Nigeria. Moreover, existing studies predominantly relied on school reports and grades, which complicates cross-regional or international comparisons of academic achievement due to significant differences in grading systems, evaluation criteria, and assessment methods across schools in different areas. A significant research gap on this topic therefore exists elsewhere in the region, including Uganda. In addition, children with SCA in East Africa differ from Nigerian children in aspects of genetics, environment, and social structure [19]. This study aimed to compare academic achievement between a sample of school-aged Ugandan children with SCA and a sample of siblings and other close family members without SCA. We hypothesized that children with SCA have poorer scores in each of the four main academic achievement areas (spelling, mathematical computation, word reading, and sentence comprehension) tested compared to the unaffected children.

## METHODS

### Study design and setting

This cross-sectional study was nested in a larger study: “Burden and Risk of Neurological and Cognitive Impairment in Paediatric Sickle Cell Anaemia in Uganda (BRAIN SAFE)” [20]. The initial BRAIN SAFE study was conducted in 2016–2018, with this additional study conducted in 2019. Study procedures and data collection were carried out at the sickle cell clinic of Uganda’s national referral hospital, Mulago Hospital Sickle Cell Clinic (MHSCC), which cares for more than 6000 patients with the disease.

### Participants

The BRAIN SAFE study had enrolled 265 children with SCA aged 1-12 years from the roster of children receiving care at MHSCC. Haemoglobin electrophoresis was used to confirm the hemoglobinopathy diagnosis of all children with SCA (HbSS or HbS-B^0^thalassemia). Seventy children, aged 6–12 years (10 in each of the seven-year age range), who had participated in the BRAIN SAFE cross-sectional study were invited to enrol in this supplemental study [20].

### Recruitment

Caregivers of 70 children with SCA ages 6-12 years who had participated in the BRAIN SAFE study were telephoned to invite their child to participate in this study. They were also asked about their willingness for an unaffected sibling child to participate as a control. For children with SCA and more than one apparently unaffected sibling within the eligible age range, only one was randomly invited to enrol from among those available. In the absence of an available age-appropriate sibling, a close relative was recruited instead. Willing parents and children were invited for informed consent (and assent for the child if 8 years and older), enrolment, and administration of study procedures. For the control participants, a blood sample was obtained for haemoglobin electrophoresis to confirm the haemoglobinopathy status of the controls (HbAA or HbAS sickle trait). Any control identified as having SCA was excluded and referred to the MHSCC for standard care. All children in either group with a history of neurological impairment before 4 months of age were excluded to reduce the risk of including those with very early adverse neurologic events likely unrelated to SCA [21].

### Sample size

The sample size was estimated based on the approximately 30% prevalence of neurocognitive abnormalities in the BRAIN SAFE SCA sample [20] that may impact academic achievement. Additionally, international studies suggest that children with SCA have an estimated three-fold increased risk of cognitive dysfunction compared to unaffected controls. [22]. Assuming a 10% prevalence of neurocognitive abnormalities in the control group (one-third of the 30% prevalence in the SCA group)[23], a minimum sample size of 63 children with SCA and 63 controls was required to achieve 80% power to detect a 20% difference in age-adjusted academic achievement z-scores between the two groups at a significance level of α = 0.05. To account for an anticipated 10% no-show rate, the adjusted sample size was set at 70 children per group.

### Clinical examination

All participants underwent standard paediatric physical examination, including anthropometry to assess height and weight, and a standardized stroke-focused neurological examination using the NIH Paediatric Stroke Scale (PedNIHSS) to assess for prior stroke [20, 24]. No child was acutely ill at the time of testing, anyone who presented to testing with an acute illness was treated and rescheduled appointments to return at a later date.

### Academic achievement testing

Age-appropriate testing for academic achievement was performed using the Wide Range Achievement Test, Fourth Edition (WRAT4). The WRAT4 is a standardized assessment tool designed to measure academic achievement for individuals ages 5 years and older across four domains: i) word reading, ii) sentence comprehension, iii) spelling, and iv) mathematical computation [25]. Two qualified psychological research assistants conducted the assessments. This assessment tool had previously been used in Uganda to assess academic achievement among children with malaria [26]. As indicated in the test’s administration manual, children scoring 4 or less on the second part of the word reading subtest, were not tested for sentence comprehension [25].

### Socio-economic status

Socioeconomic status (SES) was assessed using, an established checklist of household members, housing quality, food availability, material possessions, and water access. This SES tool has previously been validated and used among the Ugandan population [27].

### Outcome measures

The outcome measures were each of the four areas tested by the WRAT-4: spelling, math computation, word reading, and sentence comprehension, normalized for age and by sex. SCA status was the predictor variable under investigation. Covariates considered were age, sex, maternal education anaemia, and grade level. Grade level was categorised as lower primary (Grade 1-Grade 2), middle primary (Grade 3-Grade 4), and upper primary (Grade 5-Grade 6). Anaemia was defined as no anaemia ≥11g/dl, mild-to-moderate anaemia <11-8g/dl and severe anaemia, defined as <□8□g/dl (National Cancer Institute CTCAE Version 5.0, 2017) [28].

## Data analysis

Stata 16.0 was used to analyse the data, with a *p*-value <0.05 considered statistically significant [29]. The primary analysis involved the conversion of each WRAT-4 subtest total raw score to age- and sex-normalized z-scores, defined by the control group scores and were computed as:

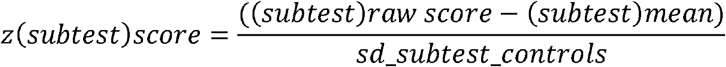

where the mean score for a child’s age and sex and standard deviation (SD) were computed by fitting a linear regression model to each outcome for all controls. Z-scores for numerical data were calculated as means with SD, with scores for categorical data calculated in percentages. We adjusted for sex, as previous studies had shown nominal sex-associated effects on academic achievement among children with SCA [30].

The Student’s t-test was used to test for differences in academic achievement between the two groups. The relationship between academic achievement, sex, maternal education and grade level was assessed using a non-parametric test (Kruskal-Wallis) and results were reported in terms of ***X***^**2**^ and the corresponding *p*-value. Spearman correlation test was used to assess the relationship between academic achievement and age. The Wilcoxon rank-sum (Mann-Whitney) test was used to assess the relationship between anaemia levels (mild-to-moderate and severe anaemia) with academic achievement.

Missing or incomplete data were present for several variables, including ‘grade level’ (no response for 52 participants, 38%), ‘anaemia’ (missing for 18 participants, 13.1%), and ‘maternal education’ (no response for 16 participants, 11.7%). The missing data were mainly attributed to participant non-responses and the omission of specific items or questions. Missing data was excluded from the analysis.

### Ethical considerations

Ethical approval for this study was provided by the Makerere University School of Medicine Research and Ethics Committee (Ref: 2016-081), Columbia University Institutional Review Board (protocol AAAR0105), and the Uganda National Council for Science and Technology (HS2114). Written consent was obtained from the parents or guardians of the study participants, with assent obtained from children aged 8 years and older.

### Patient and public involvement

Patients and the public were not involved in the design, conduct or reporting of this study. Study findings will be disseminated through open-access publication.

## RESULTS

### Socio-demographic characteristics

A total of 140 eligible children were enrolled in this study: 70 with SCA and 70 age-matched controls. Two participants with SCA were excluded for missing their appointments for testing. Among the 70 enrolled controls, one was found to have HbSS and excluded. Therefore, 137 (97.9%) of enrolled children (68 with SCA and 69 controls) underwent study procedures. As previously reported, fewer than 10% in the control group were close relatives but not siblings of the SCA sample, hence, referred to here as “sibling controls.” [31]. Among the groups with SCA and controls, 38/68 (55.9%) and 32/69 (46.4%) were male, respectively. Among the controls, 46 (66.7%) were found to have HbAA, and 23 (33.3%) had HbAS. Mean group age was 9.4±2.0 years for each group. (Table 1).

**Table 1.** Participant characteristics by sickle cell status.

| Characteristic | SCA<br>N = 68 (49.6%) |  | Control<br>N = 69 (50.4%) |  | Total N = 137<br>(100%) |  |
| --- | --- | --- | --- | --- | --- | --- |
|  | n | (%) | n | (%) | n | (%) |
| **Sex** |  |  |  |  |  |  |
| Male | 38 | (55.9) | 32 | (46.4) | 70 | (51.1) |
| **Age in years (mean, sd)** | 9.44 | (2.04) | 9.42 | (2.02) | 9.43 | (2.02) |
| **Grade Level** |  |  |  |  |  |  |
| Lower primary | 18 | (26.5) | 15 | (21.7) | 33 | (24.1) |
| Middle primary | 17 | (25.0) | 18 | (26.1) | 35 | (25.5) |
| Upper primary | 5 | (7.3) | 12 | (17.4) | 17 | (12.4) |
| No response | 28 | (41.2) | 24 | (34.8) | 52 | (38.0) |
| **Anaemia** |  |  |  |  |  |  |
| No anaemia | 0 | (0.00) | 58 | (84.1) | 58 | (42.3) |
| Mild-to-moderate<br>anaemia | 26 | (38.2) | 1 | (1.4) | 27 | (19.7) |
| Severe anaemia | 34 | (50.0) | 0 | (0.0) | 34 | (27.0) |
| Missing | 8 | (11.8) | 10 | (14.5) | 18 | (13.1) |
| **Maternal education** |  |  |  |  |  |  |
| Primary or less | 10 | (14.7) | 18 | (26.1) | 28 | (20.4) |
| Secondary | 38 | (55.9) | 34 | (49.3) | 72 | (52.5) |
| Tertiary | 11 | (16.2) | 10 | (14.5) | 21 | (15.3) |
| No response | 9 | (13.2) | 7 | (10.1) | 16 | (11.7) |
| **SES score (mean, sd)** | 7.00 | (4.0) | 7.00 | (3.0) | 7.00 | (5.0) |

\*Grade level is categorized as Lower primary (Grade 1-Grade 2), Middle primary (Grade 3-Grade 4), and Upper primary (Grade 5-Grade 6).
Mean assessed by Student's t-test

#### Hemoglobinopathy status

Children with SCA had significantly lower haemoglobin levels than the controls, as evidenced by the marked disparity (Figure 1). Detailed analysis showed all participants but one child in the control group had normal Hb levels. The one child had moderate anaemia with an Hb of 8.9 g/dl. Among the 60 children with SCA who had hb results, all had anaemia (hb<11 g/dl) with 50 (83.3%) having mild-to-moderate anaemia and the rest (16.7%) having severe anaemia.

**Figure 1.**
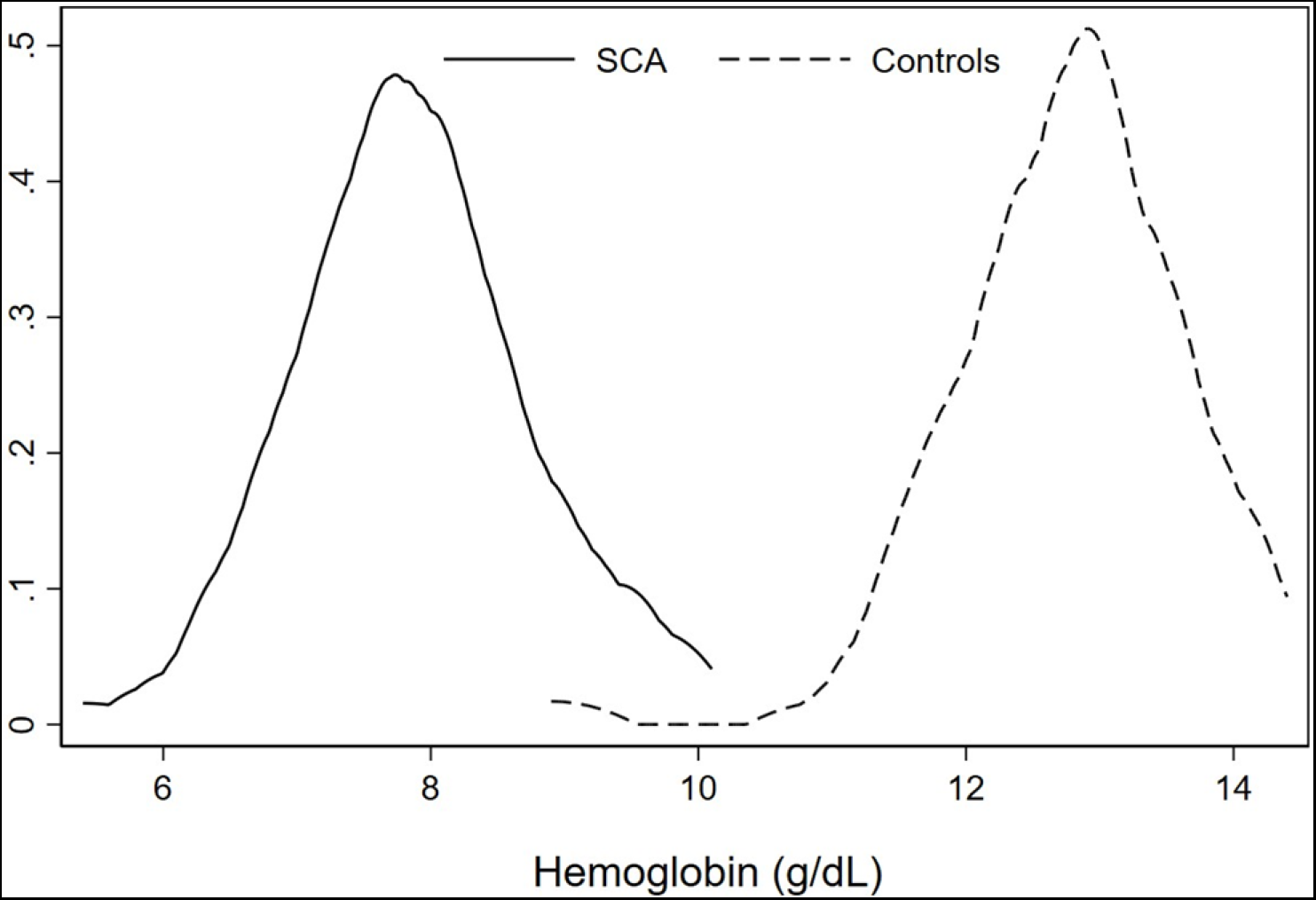
Distribution of haemoglobin in study participants The kernel plot above showed nearly perfect discrimination between children with SCA and controls based on their haemoglobin levels.

### Academic achievement

Compared to the control group, children with SCA had significantly poorer performance in spelling (mean difference [95% confidence interval] −0.36 [−0.02 to −0.69], *p*=0.04) and mathematical computation (mean difference [95% confidence interval] −0.51 [−0.17 to −0.85], *p*=0.003). No significant differences were found in performance for word reading or sentence comprehension (Table 2).

**Table 2.** Academic achievement (age- and sex-normalized Z-scores), by haemoglobinopathy status.

| Domain/Subtest | SCA (n=68)<br>Mean (SD) | Controls (n=69)<br>Mean (SD) | Difference in mean<br>(95% CI) | p-value |
| --- | --- | --- | --- | --- |
| Spelling | -0.36 (1.00) | 0.00 (0.99) | -0.36 (-0.02 to -0.69) | **0.04** |
| Mathematical computation | -0.51 (1.02) | 0.00 (0.99) | -0.51 (-0.17 to -0.85) | **0.003** |
| Word reading | -0.31 (1.20) | 0.00 (0.99) | -0.31 (0.06 to -0.68) | 0.10 |
| Sentence comprehension* | -0.07 (0.98) | 0.00 (0.98) | -0.07 (0.32 to -0.46) | 0.71 |

Age- and sex-normalized z-scores were computed using the controls from this study.
\*Children with SCA, n= 44; controls n=57. Children who scored $\leq 4$ on the second part of the word reading subtest were not administered the sentence comprehension subtest.
Student's t-test was used to compare groups.

Supplementary Table 1 demonstrates that maternal education was positively associated with spelling (*X*^*2*^ =6.62, *p*=0.04) and mathematical computation (*X*^*2*^ =8.44, *p*=0.01), while grade level is positively associated with mathematical computation (*X*^*2*^ =7.19, *p*=0.03) and sentence comprehension (*X*^*2*^ =8.00, *p*=0.02).

Assessing the sample with SCA and selected covariables (Table 3), age and spelling were found to be significant negatively correlated (ρ**=-**0.25; *p*=0.04) as further shown in Figure 2. Much as children with SCA performed poorer than the control children in mathematical computation (Table 2), this poor performance in mathematical computation among children with SCA did not vary by age (Table 3). While comparing the academic achievement performance of the sample with SCA, males performed better than their female counterparts in mathematical computation (*X*^*2*^ = 5.45, *p*=0.02).

**Table 3.**
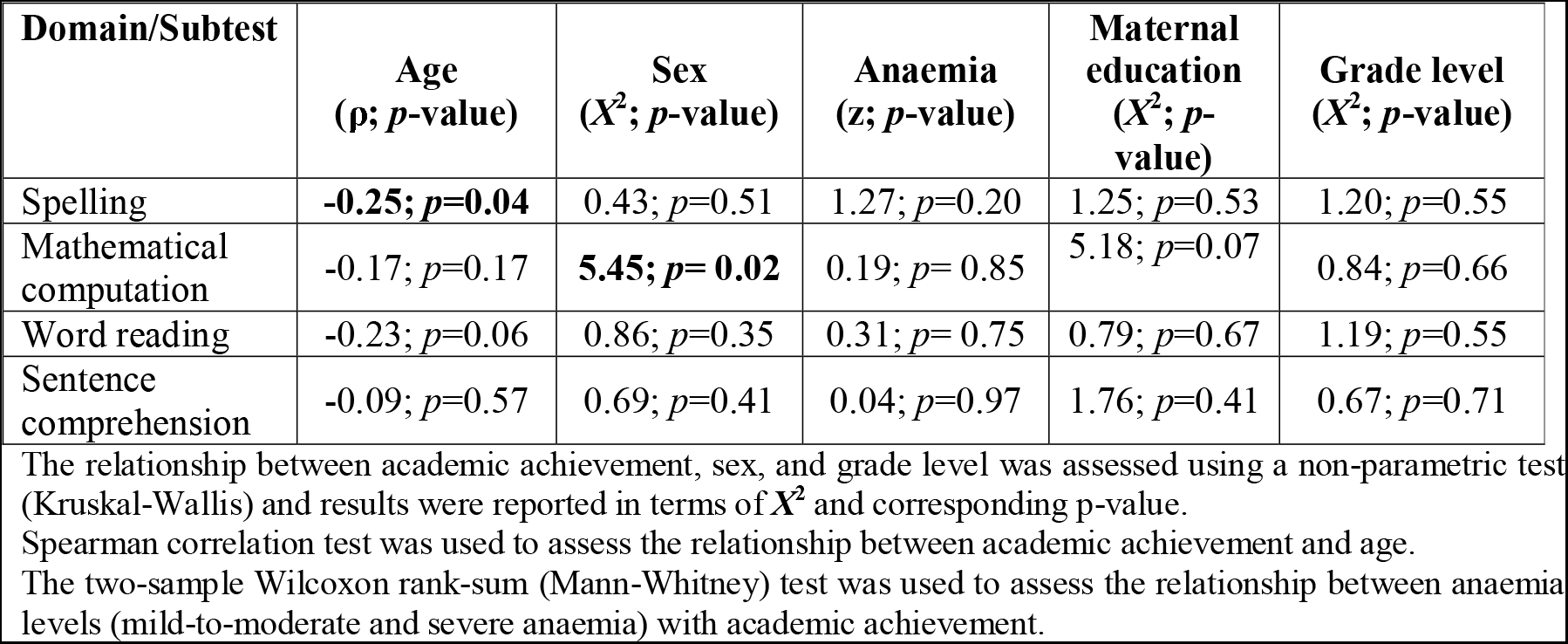
Associations between academic achievement and selected covariates in the sample with SCA.

**Figure 2.**
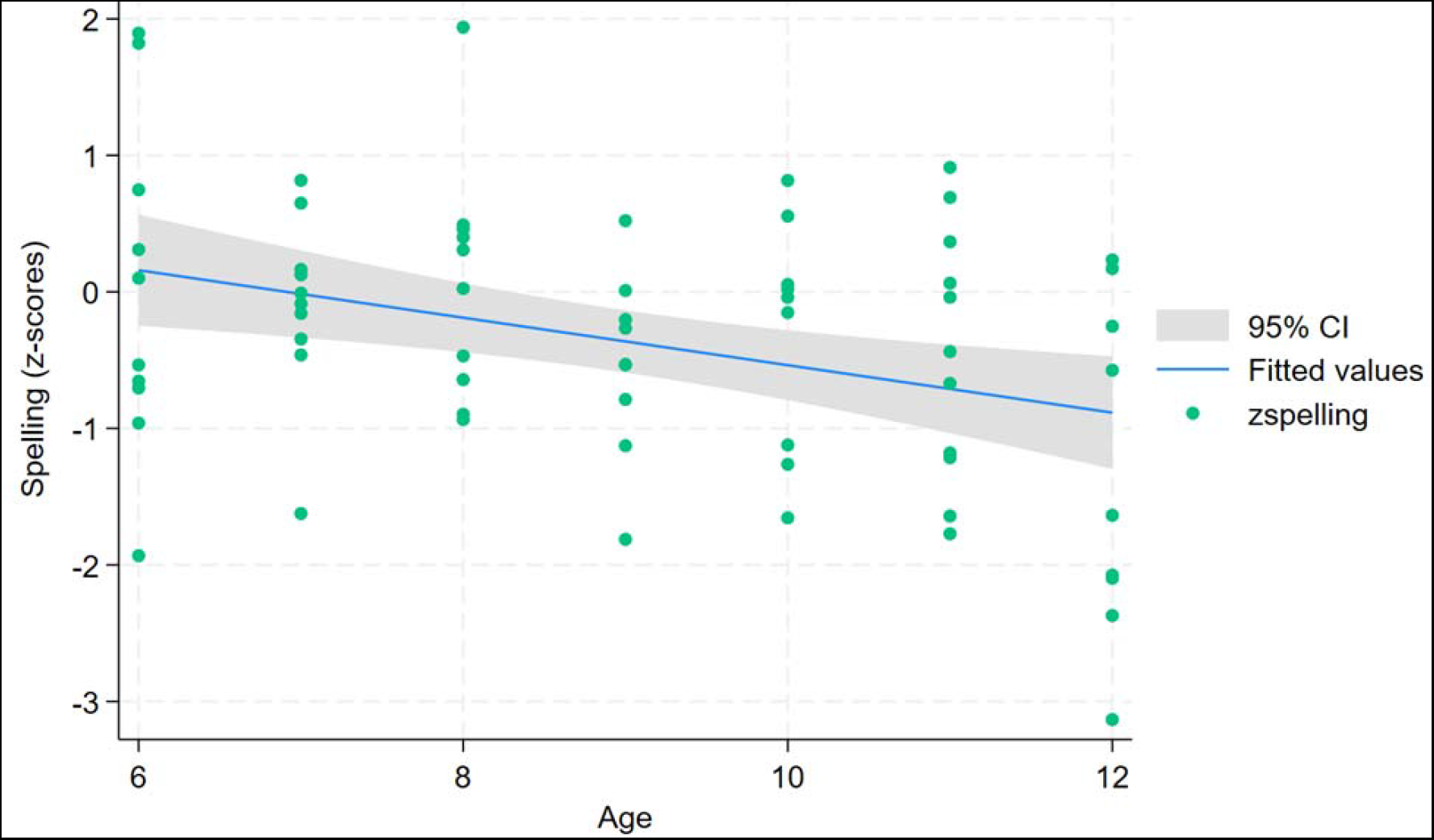
A scatter plot showing the relationship between spelling and age among children with SCA

## DISCUSSION

In this Uganda-based study, we found that a sample of children with SCA had lower academic achievement scores in the areas of spelling and math computation, but not in word reading and sentence comprehension, compared to age-matched siblings without SCA. In addition, spelling scores declined with age while girls had lower scores in math computation.

These findings add to the increasing global evidence highlighting the detrimental effect of SCA on the academic success of affected children. Our study findings of lower academic achievement, specifically poorer math and spelling scores, in children with SCA in East Africa have not been previously reported. Our findings of these specific areas of lower academic achievement are consistent with reports from high-income countries [7, 32, 33]. Children with SCA in the U.S. exhibit lower proficiency in spelling and math when assessed using the WRAT in comparison to their healthy counterparts, a trend consistent with our research findings [32, 34].

Studies in Nigeria comparing average academic scores among children aged 6 to 17 years did not show a difference in children with SCA compared to children without SCA, aligning with our observations in word reading and sentence comprehension [15, 18]. Our study of the SCA sample also suggests that age is associated with lower spelling performance. This negative age impact might be explained by increasing cognitive deficits, which in Ugandan children with SCA increased with increasing age [20]. Similar findings were described in Nigeria [17]. Biologic and/or other factors, e.g. illness-related and school absences contribute to these cumulative impacts on performance [15, 17].

Silent infarcts associated with SCA affect structural areas of working memory and attention, both of which are essential for math calculation and learning [12, 35]. Wang et al observed significantly lower math scores among U.S. children with SCA and silent infarcts detected by MRI [12]. Additionally, as children with SCA age, they can accumulate clinically “silent infarcts,” often affecting the frontal and parietal lobes which are areas crucial for language processing [11, 17, 36-38]. We previously identified that a large proportion of the BRAIN SAFE participants had clinically “silent” cerebral infarcts [39]. The presence or location of MRI-identified lesions in specific brain regions and their impact on academic achievement were not studied in our current study.

Girls with SCA performed poorer in math computation compared to their age-normalized male counterparts. Girls in Uganda have been reported to perform less favourably in mathematics compared to boys [40]. Similar patterns have been observed in other African countries, suggesting a broader regional trend [41]. In contrast, a Jamaican study of children with SCA reported poorer academic achievement among boys between the ages of 11 and 13 years [30]. These findings suggest that sex differences in academic performance among children with SCA may vary across different regions and ages due to the effects of SCA and possibly quality of education and/or additional variables.

Limited knowledge exists regarding the relationship between grade level and academic achievement in children with SCA within African studies. Despite advancing to higher grade levels, children with SCA in our study maintained consistent academic performance, diverging from the anticipated trend of better performance as grade level increases observed in the control group [42, 43]. This finding is consistent to prior research supporting the conclusion that educational attainment among adult individuals with SCA does not necessarily translate to better performance or mental processing [44].

### Study limitations

Our study was unable to collect data on grade repetition, school absenteeism, or school quality due to the many different schools attended by this sample. MRI results were not included in this report, as only a selected subset of the children had undergone imaging, and imaging was as much as nine months prior to this testing. Per WRAT-4 scoring standards, children who scored <4 on the second part of the word reading subtest were not administered the sentence comprehension subtest. Despite these limitations, our study employed a rigorous methodological approach using a robust standardized assessment of academic achievement.

## Conclusion

To our knowledge, this study provides the first evidence of lower academic achievement among children with SCA in East Africa. Our findings suggest the presence of academic disadvantages for children with SCA in Uganda compared to unaffected siblings. Academic performance can have long-term effects on grade attainment and employment opportunities, thereby highlighting the importance of this topic. Overall, our study suggests that children with SCA in the region may need educational assessment and additional educational support for academic achievement, especially in mathematics and spelling subjects [45]. Our findings also suggest a need for targeted interventions to address the unique challenges faced by children with SCA in achieving academic success. Future studies with brain imaging may add neuro-anatomical correlates of SCA that significantly impact academic achievement and also evaluate potential sex disparities in academic achievement among children with SCA.

## Data Availability

All data produced in the present study are available upon reasonable request to the authors

## Acknowledgements

We acknowledge and thank the support from Global Health Uganda and the caregivers, patients, research assistants and staff at Mulago Hospital Sickle Cell Clinic for their central participation in the study.

## Author Contributions

SKN: Conceptualization, Formal analysis, Investigation, Methodology, Supervision, Writing and editing. PB: Conceptualization, Investigation, Methodology, Project administration, Resources, Supervision, Writing – review and editing. RO: Conceptualization, Investigation, Methodology, Project administration, Resources, Supervision, Writing – review and editing. OS: Formal analysis, Writing – review & editing. BN: Data curation, Review and editing. AB: Data curation, Review and editing. GN: Data curation, Review and editing. MK: Data curation, Project administration, Writing – review and editing. AJN: Methodology, Review and editing. DK: Methodology, Formal analysis. DM: Conceptualization, Supervision, Writing – review and editing. PK: Resources, Supervision, Writing – review and editing. EM: Conceptualization, Methodology, Resources, Supervision, Writing – review and editing. JMS: Methodology, Formal analysis, Writing – review and editing. NSG: Conceptualization, Funding acquisition, Investigation, Supervision, Writing – original draft, Writing – review & editing. RI: Conceptualization, Investigation, Project administration, Resources, Supervision, Writing – original draft, Writing – review & editing.

## Funding

This work was supported by the National Institute of Health (NIH) and Fogarty International Centre (FIC) under four grant awards; 1R21HD089791, 3R01HD096559-04, R01HD096559 (MPIs Idro, Green) and D43TW010928 (MPIs John, Idro). The content of this report is solely the responsibility of the authors and does not represent the official views of NIH. The funding source had no role in the design of the study and collection, analysis, and interpretation of data and in writing the manuscript.

## Competing interests

The authors have no competing interests.

## Patient consent for publication

Not applicable.

## Ethical approval

The study was approved by the Makerere University School of Medicine Research and Ethics Committee and the Columbia University Institutional Review Board.

## Data sharing statement

Data are available upon reasonable request.

## Notes

### Competing Interest Statement

The authors have declared no competing interest.

### Funding Statement

This study was funded by the National Institute of Health (NIH) and Fogarty International Centre (FIC) under four grant awards; 1R21HD089791, 3R01HD096559-04, R01HD096559 (MPIs Idro, Green) and D43TW010928 (MPIs John, Idro).

### Author Declarations

Makerere University School of Medicine Research and Ethics Committee of Makerere University and Columbia University Institutional Review Board of Columbia University gave ethical approval for this work

