## Supplemental Table 1 for "Academic achievement in Ugandan children with sickle cell anaemia: A cross-sectional study"

Supplementary Table 1. Correlations between academic achievement and selected covariates in the control group

| **Domain/Subtest** | **Maternal Education**  **(*X*^2^; p-value)** | **Grade level**  **(*****X*^2^; p-value)** |
| --- | --- | --- |
| Spelling | **6.62; p=0.04** | 5.66; p=0.06 |
| Mathematical computation | **8.44; p= 0.01** | **7.19; p=0.03** |
| Word reading | 2.97; p=0.23 | 0.81; p=0.67 |
| Sentence comprehension | 5.78; p=0.06 | **8.00; p=0.02** |

The relationship between academic achievement and covariates were assessed using non-parametric test (Kruskal-Wallis) and results reported in terms of ***X*^2^** and corresponding p-value.
